# Excessive matrix metalloproteinase-1 and hyperactivation of endothelial cells occurred in COVID-19 patients and were associated with the severity of COVID-19

**DOI:** 10.1101/2021.01.19.21250115

**Authors:** Fahim Syed, Wei Li, Ryan F. Relich, Patrick M. Russell, Shanxiang Zhang, Michelle K. Zimmerman, Qigui Yu

## Abstract

COVID-19 starts as a respiratory disease that can progress to pneumonia, severe acute respiratory syndrome (SARS), and multi-organ failure. Growing evidence suggests that COVID-19 is a systemic illness that primarily injures the vascular endothelium, yet the underlying mechanisms remain unknown. SARS-CoV-2 infection is believed to trigger a cytokine storm that plays a critical role in the pathogenesis of endothelialitis and vascular injury, eventually leading to respiratory and multi-organ failure in COVID-19 patients. We used a multiplex immunoassay to systematically profile and compare 65 inflammatory cytokines/chemokines/growth factors in plasma samples from 24 hospitalized (severe/critical) COVID-19 patients, 14 mild/moderate cases, and 13 healthy controls (HCs). Patients with severe/critical and mild/moderate COVID-19 had significantly higher plasma levels of 20 analytes than HCs. Surprisingly, only one cytokine (MIF) was among these altered analytes, while the rest were chemokines and growth factors. In addition, only MMP-1 and VEGF-A were significantly elevated in hospitalized COVID-19 patients when compared to mild/moderate cases. Given that excessive MMP-1 plays a central role in tissue destruction in a wide variety of vascular diseases and that elevated VEGF-A, an EC activation marker, increases vascular permeability, we further studied MMP-1 enzymatic activity and other EC activation markers such as soluble forms of CD146, ICAM-1, and VCAM-1. We found that plasma MMP-1 enzymatic activity and plasma levels of MMP-1 and EC activation markers were highly dysregulated in COVID-19 patients. Some dysregulations were associated with patients’ age or gender, but not with race. Our results demonstrate that COVID-19 patients have distinct inflammatory profiles that are distinguished from the cytokine storms in other human diseases. Excessive MMP-1 and hyperactivation of ECs occur in COVID-19 patients and are associated with the severity of COVID-19.

## Introduction

Coronavirus disease 2019 (COVID-19), an infectious disease caused by a novel coronavirus (severe acute respiratory syndrome coronavirus 2 or SARS-CoV-2), has created an unprecedented global health and economic crisis. By the end of 2020, confirmed COVID-19 cases surpassed 84 million globally, resulting in over 1.8 million deaths. COVID-19 patients can experience a range of manifestations, from being asymptomatic, to having mild, moderate, severe symptoms, to having a critical illness. Growing evidence shows that COVID-19 is a vascular illness, not solely a respiratory disease. Histopathological examinations of post-mortem tissues of COVID-19 patients have revealed ***(1)*** diffuse alveolar damage with perivascular infiltration of inflammatory cells^1-5^, ***(2)*** extensive damage to the lining of blood vessels throughout the body^6-8^, ***(3)*** severe endothelial injury and widespread thrombosis in the lungs, heart, liver, kidney, and small intestine^1,5-8^, ***(4)*** viral particles in endothelial cells (ECs) of the glomerular capillary loops^6^, and ***(5)*** caspase-3-positive apoptotic ECs in the lung and intestine tissues^6^. In addition, a recent study has shown that COVID-19 patients in intensive care units (ICUs) have higher counts of circulating ECs (CECs) than non-ICU patients^9^. CECs are stressed cells detached from injured blood vessels, thereby indicating severe vascular injury^10^.

Collectively, patients with severe COVID-19 exhibit impaired endothelial and microcirculatory functions across vascular beds of different organs, which may be particularly relevant for vulnerable individuals with pre-existing endothelial dysfunctions such as diabetes, obesity, hypertension, and cardiovascular diseases, all of which are associated with adverse outcomes in COVID-19^6,11-14^.

The pathological mechanisms underlying vascular injury in COVID-19 remain unclear, although cytokine storm syndrome (CSS) and direct SARS-CoV-2 infection are considered contributors. SARS-CoV-2 is a member of the family *Coronaviridae*, genus *Betacoronavirus*, and is closely related to SARS-CoV that caused the 2003 SARS pandemic^15-17^. CSS plays a critical role in the pathogenesis of SARS-CoV infection and represents a major cause of morbidity in SARS patients^18,19^. Elevated circulating concentrations of IL-6, IFN-γ, IL-8, IL-18, TGF-β, IP-10, MCP-1, and MIG have been reported in SARS patients when compared with healthy controls (HCs)^20,21^. Circulating IL-6 levels are also elevated in COVID-19 patients and are cited as evidence of COVID-19 CCS^22,23^. However, the levels of IL-6 and other inflammatory cytokines such as IL-8 are significantly less elevated in patients with critical COVID-19 than the values typically reported in patients with CSS such as septic shock with and without acute respiratory distress syndrome (ARDS)^24^. Indeed, the concentrations of IL-6, IL-8, and TNF-α, three of the most important inflammatory mediators in human diseases with CSS^25^, in COVID-19 patients are similar to those found in ICU patients with cardiac arrest or trauma, conditions that are not notable for cytokine storms^24^. These findings bring into question whether a cytokine storm occurs in COVID-19, and whether IL-6, IL-8, and TNF-α act as key inflammatory mediators for fatal manifestations in patients with severe or critical COVID-19^25^. Our results echo recent NIH guidelines that note there are insufficient data to recommend IL-6 inhibitors for the treatment of COVID-19^26^. In fact, efforts to combat cytokine storm in patients with severe COVID-19 have proven unsuccessful^27^.

SARS-CoV-2 uses angiotensin-converting enzyme 2 (ACE2) as a primary receptor for viral binding and entry^28-32^. Viral entry is also facilitated by transmembrane protease serine 2 (TMPRSS2) and neuropilin-1 (NRP1)^31,33,34^. ACE2, TMPRSS2, and NRP1 are highly abundant on the lung alveolar type II epithelial (AT2) cells, rendering AT2 cells highly susceptible to productive SARS-CoV-2 infection^33,35-37^. ACE2, TMPRSS2, and NRP1 are also expressed on the surface of ECs^33,35,38-40^, albeit at lower levels than in AT2 cells^33^, suggesting that these molecules may facilitate direct SARS-CoV-2 infection of ECs. Indeed, SARS-CoV-2 particles were observed in ECs in kidney tissues from COVID-19 patients^8^. However, a recent unreferenced preprint reported that ECs derived from human peripheral blood mononuclear cells (PBMCs), human lung microvascular ECs, and human aortic ECs did not have detectable mRNA of ACE2 and TMPRSS2 and were resistant to SARS-CoV-2 infection *in vitro*^41^. Although this study shows results that are not consistent with both their own report^42^ and previous studies^33,35,38-40^, it highlights the need for further research on the pathogenic mechanisms underlying vascular injury in COVID-19.

To explore the profile and key players of the cytokine storm in COVID-19, we used a multiplex immunoassay to simultaneously measure 65 inflammatory cytokines/chemokines/growth factors in plasma samples from hospitalized (severe/critical) COVID-19 patients, mild/moderate cases, and HCs. Surprisingly, matrix metalloproteinase-1 (MMP-1) and vascular endothelial growth factor A (VEGF-A), not conventional inflammatory cytokines such as IL-6, were two of the most unambiguously elevated inflammatory factors in hospitalized COVID-19 patients when compared to mild/moderate cases or HCs. Spearman correlation analysis revealed that the plasma levels of MMP-1 and VEGF-A in hospitalized COVID-19 patients were positively correlated, suggesting that there may be an interaction between MMP-1 and VEGF-A in COVID-19. Given that excessive MMP-1 plays a central role in tissue destruction in a wide variety of vascular diseases and that elevated VEGF-A, an EC activation markers, increases vascular permeability^43^, we further studied MMP-1 enzymatic activity and other EC activation markers including soluble forms of CD146, intercellular adhesion molecule 1 (ICAM-1), and vascular cell adhesion molecule 1 (VCAM-1) in our cohort of COVID-19 patients and HCs. We performed correlation analysis among plasma levels of MMP-1, EC activation markers, and inflammatory cytokines/chemokines, MMP-1 enzymatic activity, and patients’ demographics and clinical parameters. We found that excessive MMP-1 and hyperactivation of ECs occurred in COVID-19 patients and were correlated with the severity of COVID-19.

## Materials and methods

### Study subjects and ethical considerations

This study was performed with the approval of the Institutional Review Boards at Indiana University School of Medicine. Blood samples were drawn after each participant provided a written informed consent form.

Combined nasopharyngeal-oropharyngeal swab specimens collected from study subjects were tested using either the cobas^®^ SARS-CoV-2 Test (Roche Diagnostics, Indianapolis, IN) real-time PCR assay performed on either the cobas^®^ 6800 or 8800 platforms or the NxTAG^®^ CoV Extended Panel (Luminex Molecular Diagnostics, Toronto, ON, Canada) reverse-transcription PCR assay. Diagnostic tests were used in accordance with manufacturer instructions for use pursuant to U.S. Food and Drug Administration Emergency Use Authorization.

The study subjects included 24 hospitalized COVID-19 patients, 14 mild/moderate cases, and 13 HCs. Hospitalized COVID-19 patients included SARS-CoV-2-infected individuals who developed severe illness (dyspnea, hypoxia, >50% lung involvement on imaging, or required oxygen support^44^) or critical disease with complications such as respiratory failure, disseminated intravascular coagulation (DIC), and/or multi-organ failure^44^. Some of these hospitalized patients were admitted to ICUs. Patients with mild/moderate COVID-19 were characterized with mild respiratory symptoms (nasal congestion, runny nose, and a sore throat) or mild pneumonia^44^. All COVID-19 patients were treated in Indiana University (IU) Health hospitals in Indianapolis, Indiana, during May – December 2020. Plasma samples from HCs were selected from our banked blood samples that were collected before the COVID-19 pandemic as described in our previous reports^45,46^. Demographics of HCs were matched with COVID-19 patients so that there were no significant differences between age or gender of HCs and COVID-19 subjects. The demographic and clinical characteristics of COVID-19 patients and HC demographics were summarized in Table 1.

**Table 1.** Demographics and clinical characteristics of COVID-19 patients vs HCs.

|  | HC (n=13) | Mild (n=14) | Hosp (n=24) |
| --- | --- | --- | --- |
| Age (years) | 56.6 (42.1 - 65.5) | 59.0 (43.0 - 63.0) | 64.0 (48.5 - 73.0) |
| Gender (male) | 6 (46.3%) | 6 (42.9%) | 10 (41.7%) |
| Race |  |  |  |
| African American | 5 (38.5%) | 7 (50.0%) | 10 (41.7%) |
| White | 7 (53.8%) | 6 (42.9%) | 13 (54.2%) |
| Other | 1 (7.7%) | 1 (7.1%) | 1 (4.1%) |
| D-Dimer (ng/mL) | <250 | 282 (259 - 290) | 527 (244 - 814)*** |
| CRP (mg/L) | <10 | 126 (89 - 268)* | 830 (642 - 1,273)*** |
| Ferritin (ng/mL) | <300 | 350 (145 - 939) | 311 (88 - 1,078) |
| WBC (cell number/μL) | 4,000-11,000 | 5,450 (4,200 - 8,100) | 8,000 (5,800 - 12,150) |
| Neutrophil (ANC/μL) | 1,500 - 8,000 | 3,600 (2,550 - 4,800) | 6,300 (4,400 - 10,300) |
| Lymphocyte (ALC/μL) | 1,000-4,800 | 1,100 (650 - 1,850) | 610 (522 - 660)# |

**Note:** Values of D-Dimer, CRP, ferritin, WBC, ANC, and ALC in HCs are the reference numbers for healthy population. \* $p<0.05$ , \*\*\* $p<0.001$ (increased), & # $p<0.05$ (decreased): comparison between mild/moderate or hospitalized COVID-19 and HCs. WBC, white blood cell count; ANC, absolute neutrophil count; ALC, absolute lymphocyte count; CRP, c-reactive protein; HC, healthy control; Mild, patients with mild/moderate COVID-19; Hosp, hospitalized COVID-19 patients.

### Multiplex and ELISA immunoassays

Peripheral blood was collected in heparin-coated BD Vacutainer Blood Collection tubes (BD Biosciences, Franklin Lakes, NJ). Blood samples were centrifuged within 12 h of collection at 700 g for 20 min at room temperature without brake. The top layer (plasma) was harvested and stored at -80 °C until use. Plasma concentrations of 65 human cytokines/chemokines/growth factors including 33 cytokines (G-CSF, GM-CSF, IFN-α, IFN-γ, IL-1α, IL-1β, IL-2, IL-3, IL-4, IL-5, IL-6, IL-7, IL-8, IL-9, IL-10, IL-12p70, IL-13, IL-15, IL-16, IL-17A, IL-18, IL-20, IL-21, IL-22, IL-23, IL-27, IL-31, LIF, M-CSF, MIF, TNF-α, TNF-β, and TSLP), 18 chemokines (CXCL13, CXCL5, CCL11, CCL24, CCL26, CX3CL1, CXCL1, CXCL10, CXCL11, MCP-1, MCP-2, MCP-3, MDC, MIG, MIP-1α, MIP-1β, MIP-3α, and SDF-1α), 6 growth factors/regulators (FGF-2, HGF, MMP-1, NGF-β, SCF, VEGF-A), and 8 soluble receptors (APRIL, BAFF, CD30, CD40L, IL-2R, TNF-RII, TRAIL, and TWEAK) were simultaneously measured using a magnetic bead-based multiplex kit (65-Plex Human ProcartaPlex^™^ Panel, EPX650-10065-901, Invitrogen, Carlsbad, CA) according to the manufacturer’s instructions. Briefly, plasma samples were incubated with beads at room temperature for 2 h with shaking at 500 rpm, and subsequently washed twice using a magnetic plate washer (eBioscience, San Diego, CA). The beads were incubated with the biotinylated antibodies for 1 h at room temperature with shaking at 500 rpm. After washing, beads were incubated with streptavidin-PE for 30 min at room temperature with shaking at 500 rpm. After washing twice, beads were resuspended in PBS and read on a BioPlex 200 system (Bio-Rad, Hercules, CA) with a setting of 40 beads per bead set and 150 seconds per well. The standards at four-fold serial dilutions were run on each plate in duplicate and used to calculate the concentrations of cytokines/chemokines/growth factors using the Bio-Plex Manager Software (Bio-Rad, Hercules, CA).

Plasma levels of soluble CD146 (sCD146), soluble ICAM-1 (sICAM-1), soluble VCAM-1 (sVACM-1), and intestinal fatty-acid binding protein (I-FABP) were quantified using the Human CD146 DuoSet ELISA Kit, the Human ICAM-1 DuoSet ELISA Kit, the Human VCAM-1 DuoSet ELISA Kit, and the Human I-FABP DuoSet ELISA kit (all from R&D Systems, Minneapolis, MN), respectively, according to manufacturer’s instructions. ELISA results were recorded using a microplate reader system (Bio-Tek, Winooski, VT).

### Quantitative determination of human active MMP-1 in plasma samples

Enzymatic activity of plasma MMP-1 was determined using the Human Active MMP-1 Fluorokine E kit (F1M00, R&D Systems, Minneapolis, MN) as per manufacturer’s instructions. Briefly, diluted plasma samples and MMP-1 standards were added to the wells that were pre-coated with a monoclonal antibody specific for human MMP-1. After washing to remove unbound substances, amino-phenyl mercuric acetate (APMA), an activation reagent of MMP-1, was added to the standards, but not the plasma samples. After washing, a fluorogenic substrate linked to a quencher molecule was added and any active enzyme present would cleave the peptide linker between the fluorophore and the quencher molecule, generating a fluorescent signal that is proportional to the amount of enzyme activity in an individual sample. Thus, plasma levels of active MMP-1 were quantitatively detected using a Synergy H1 Hybrid Multi-Mode Reader (BioTek, Winooski, VT), where fluorescence emission was recorded in relative fluorescence unit (RFU).

### Statistical analysis

Statistical analysis was performed using GraphPad Prism 6.0 (La Jolla, CA). Data were expressed as mean ± standard error of the mean (SEM) or mean ± standard deviation (SD) unless otherwise indicated. All data were tested for suitability for parametric or non-parametric analysis. Differences between two groups were calculated using the Mann Whitney test.

Kruskal-Wallis test with Dunn’s corrections was used for comparisons among three groups. Chi-square test was used for comparison between groups for categorical variables. The linear relationship between two variables was analyzed using the Spearman correlation test. *P* <0.05 was considered statistically significant.

## Results

### Characteristics of study subjects

The study subjects included 24 hospitalized patients with severe/critical COVID-19, 14 patients with mild/moderate COVID-19, and 13 HCs. Severe/critical COVID-19 patients were hospitalized in Indiana University (IU) Health hospitals in Indianapolis, Indiana and mild/moderate COVID-19 patients visited IU Health hospitals between May – December 2020. HCs were recruited before the COVID-19 pandemic for our research on immunopathogenesis of HIV infection and alcoholic hepatitis as described in our previous reports^45-48^. The demographics and clinical characteristics of these subjects were summarized in Table 1. There were no differences in age, gender, and race distributions among these three groups. Hospitalized COVID-19 patients had lower absolute lymphocyte count (ALC), higher erythrocyte sedimentation rate (ESR), and higher levels of D-dimer, C-reactive protein (CRP), and ferritin than patients with mild/moderate COVID-19 (Table 1). There were no differences in white blood cell (WBC) or neutrophil counts between the hospitalized patients and patients with mild/moderate COVID (Table 1).

### Profiles of inflammatory cytokines/chemokines/growth factors in hospitalized COVID-19 patients, mild/moderate cases, and HCs

COVID-19 patients present varied clinical features, ranging from asymptomatic, mild/moderate, severe, to critical illness. The pathogenesis of severe or critical COVID-19 is complex and has been suggested to include a cytokine storm that sustains an aberrant systemic immune response^49^. To elucidate the profile and key pathogenic inflammatory mediators of the cytokine storm in COVID-19 patients, we used a multiplex immunoassay to simultaneously detect plasma levels of 65 cytokines/chemokines/growth factors in hospitalized (severe/critical) patients, mild/moderate cases, and HCs. In comparison to HCs, hospitalized and mild/moderate COVID-19 patients had significantly higher plasma levels of 20 of these analytes (Table 2).

**Table 2.** Plasma levels of cytokines/chemokines/growth factors in COVID-19 and HC.

| Analyte | HC (n=13) | Mild (n=14) | Hosp (n=24) |
| --- | --- | --- | --- |
| BLC (CXCL13) | 36 (17 - 73) | 92 (45 - 183)* | 107 (75 - 185)*** |
| ENA-78 (CXCL5) | 22.8 (4.6 - 32.4) | 94.5 (68.0 - 69.0)*** | 108.1 (66.6 - 178.2)*** |
| Eotaxin-3 (CCL26) | 2.2 (2.2 - 2.2) | 6.0 (2.2 - 20.7)** | 9.6 (6.3 - 18.9)*** |
| Fractalkin (CX3CL1) | 2.5 (1.7 - 2.5) | 3.9 (2.8 - 5.2)* | 3.6 (2.7 - 5.6)*** |
| IP-10 (CXCL10) | 6.4 (3.1 - 9.7) | 29.1 (9.2 - 69.9)** | 28.8 (16.7 - 87.4)*** |
| I-TAC (CXCL11) | 11.1 (11.1 - 35.8) | 165.6 (92.6 (287.1)** | 196.8 (114.2 - 491.2)*** |
| MIG (CXCL9) | 8.6 (8.6 - 10.3) | 37.8 (27.4 - 69.1)*** | 26.2 (12.7 - 53.2)** |
| MIP-1α (CCL3) | 5.3 (3.1 - 26.3) | 10.0 (3.1 - 19.2)*** | 9.4 (3.5 - 19.4)*** |
| MIP-1β (CCL4) | 4.6 (4.6 - 4.6) | 20.0 (10.0 - 45.8)*** | 19.6 (8.8 - 32.7)*** |
| SDF-1α (CXCL12) | 679 (472 - 989) | 1099 (815 - 1274)* | 1087 (718 - 1275)** |
| **HGF** | **11.9 (6.4 - 17.9)** | **54.8 (28.4 - 141.1)\*** | **163.2 (77.1 - 357.3)\*\*\*** |
| **MMP-1** | **73.8 (30.9 - 96.1)** | **316.8 (189.0 - 589.5)\*\*** | **863.4 (551.4 - 1880)\*\*\*,#** |
| **SCF** | **4.6 (3.4 - 8.6)** | **15.4 (12.4 - 43.6)\*\*** | **16.6 (8.3 - 26.2)\*\*** |
| **VEGF-A** | **6.8 (6.8 - 8.2)** | **124.5 (86.0 - 293.0)\*\*\*** | **353.9 (235.1 - 701.7)\*\*\*,#** |
| APRIL | 745 (602 - 913) | 1951 (1236 - 4852)*** | 1883 (1239 - 2515)*** |
| CD30 | 100 (38 - 126) | 409 (194 - 936)*** | 274 (178 - 703)*** |
| IL-2R (CD25) | 84 (84 - 1414) | 6474 (2006 - 9316)** | 6884 (4217 - 9539)*** |
| TNF-RII | 78.6 (59.1 - 96.4) | 221.2 (158.7 - 262.3)*** | 202.2 (150.2 - 256.1)*** |
| TRAIL (CD253) | 11.0 (11.0 - 29.5) | 63.3 (41.9 - 103.3)** | 41.4 (24.5 - 415.2)* |
| **MIF** | **37.7 (19.7 - 48.4)** | **90.7 (80.3 - 125.1)\*\*\*** | **89.5 (70.7 - 157.2)\*\*\*** |
| MCP-1 (CCL2) | 41.1 (21.8 - 59.5) | 109.4 (40.1 - 281.5) | 172.2 (84.2 - 363.4)** |
| MCP-2 (CCL8) | 2.8 (0.8 - 4.9) | 4.4 (3.1 - 6.2) | 5.0 (2.6 - 8.6)* |
| IL-6 | 7.9 (7.9 - 7.9) | 7.9 (7.9 - 7.9) | 41.9 (7.9 - 76.1)* |
| IL-18 | 9.7 (4.2 - 9.7) | 9.7 (9.1 - 20.5) | 21.8 (9.7 - 56.5)*** |
| CD40L (CD154) | 4.8 (4.8 - 82.0) | 29.5 (17.6 - 71.2) | 36.6 (19.7 - 59.7) |
| Eotaxin (CCL11) | 4.2 (21.9 - 99.0) | 54.8 (17.8 - 103.0) | 51.2 (31.5 - 128.7) |
| G-CSF | 9.9 (9.9 - 33.8) | 16.5 (9.9 - 36.4) | 28.8 (13.7 - 38.9) |
| GM-CSF | 15.9 (15.9 - 54.0) | 15.9 (15.9 - 15.9) | 15.9 (15.9 - 15.9) |
| IL-2 | 36.3 (22.0 - 104.1) | 42.4 (22.0 - 106.7) | 53.9 (22.0 - 78.8) |
| IL-10 | 2.2 (2.2 - 2.9) | 1.8 (0.9 - 3.2) | 2.3 (1.3 - 4.4) |
| IL-16 | 799 (412- 1175) | 1091 (865 - 1362) | 936 (736 - 1386) |
| IL-17A | 24.3 (24.3 - 358.2) | 24.3 (24.3 - 180.3) | 24.3 (24.3 - 187.0) |
| IL-20 | 9.1 (7.6 - 50.2) | 9.1 (6.1 - 31.7) | 8.8 (2.0 - 21.2) |
| IL-21 | 9.1 (9.1 - 9.1) | 9.6 (8.9 - 33.8) | 13.8 (7.3 - 27. 2) |
| IFN-γ | 13.1 (6.4 - 13.1) | 11.2 (7.3 - 15.5) | 18.3 (8.9 - 16.4) |
| MDC (CCL22) | 46.2 (20.1 - 74.5) | 104.6 (51.9 - 164.8) | 62.9 (20.0 - 119.3) |
| MIP-3α | 21.0 (21.0 - 21.0) | 24.5 (15.2 - 57.9) | 44.8 (16.8 - 96.4) |
| TSLP | 4.7 (2.4 - 11.6) | 4.6 (2.3 - 13.2) | 6.4 (4.0 - 8.8) |
| TWEAK | 409 (216 - 1859) | 935 (469 - 1867) | 799 (566 - 1566) |
| IL-15 | 9.0 (3.3 - 27.9) | 3.3 (3.3 - 3.7) | 3.3 (3.3 - 19.4) |
| IL-21 | 9.1 (9.1 - 9.1) | 9.6 (8.9 - 33.8) | 13.8 (7.3 - 27.2) |
| IL-22 | 17.8 (17.8 - 17.8) | 17.8 (17.8 - 34.5) | 17.8 (17.8 - 23.6) |
| IL-27 | 17.4 (17.4 - 17.4) | 17.4 (17.4 - 22.8) | 17.4 (17.4 - 17.4) |
| *bNGF* | 0/13 | 1/14 | 1/24 |
| *Eotaxin-2 (CCL24)* | 0/13 | 2/14 | 4/24 |
| *FGF-2* | 0/13 | 0/14 | 1/24 |
| *Gro-α (CXCL1)* | 0/13 | 0/14 | 1/24 |
| *IFN-α* | 1/13 | 1/14 | 4/24 |
| *IL-1α* | 0/13 | 1/14 | 2/24 |
| *IL-1β* | 3/13 | 3/14 | 6/24 |
| *IL-3* | 1/13 | 5/14 | 8/24 |
| *IL-4* | 0/13 | 0/14 | 2/24 |
| *IL-5* | 1/13 | 1/14 | 2/24 |
| *IL-7* | *0/13* | *0/14* | *0/24* |
| *IL-8* | *1/13* | *1/14* | *4/24* |
| *IL-9* | *0/13* | *1/14* | *1/24* |
| *IL-12p70* | *0/13* | *1/14* | *0/24* |
| *IL-13* | *0/13* | *1/14* | *1/24* |
| *IL-23* | *0/13* | *0/13* | *1/24* |
| *IL-31* | *0/13* | *1/14* | *1/24* |
| *LIF* | *0/13* | *0/14* | *1/24* |
| *MCP-3 (CCL7)* | *0/13* | *1/14* | *0/24* |
| *M-CSF* | *0/13* | *0/14* | *0/24* |
| *TNF-α* | *0/13* | *0/14* | *0/24* |
| *TNF-β* | *0/13* | *0/14* | *0/24* |

**Note:** Data are expressed as median (interquartile range or IQR). HC, healthy control; Mild, patients with mild/moderate COVID-19; Hosp, hospitalized COVID-19 patients. Kruskal-Wallis test with Dunn's corrections was used for comparisons among three groups of HCs, mild/severe cases, and hospitalized COVID-19 patients. \* $p < 0.05$ , \*\* $p < 0.01$ , or \*\*\* $p < 0.001$ for comparison between hospitalized COVID-19 and HCs, # $p < 0.05$ for comparison between hospitalized COVID-19 patients and mild/moderate cases.

Among these altered analytes, macrophage migration inhibitory factor (MIF) was the only inflammatory cytokine, all others were chemokines (BLC/CXCL13, ENA-78/CXCL5, Eotaxin-3/CCL26, Fractalkine/CX3CL1, IP-10/CXCL10, I-TAC/CXCL11, MIG/CXCL9, MIP-1α/CCL3, MIP-1β/CCL4, and SDF-1α/CXCL12), growth factors (HGF, MMP-1, SCF, and VEGF-A), and soluble receptors (APRIL, CD30, IL-2R/CD25, TNF-RII, and TRAIL/CD253) (Table 2).

Surprisingly, MMP-1 and VEGF-A were the only two elevated inflammatory factors that were significantly higher in hospitalized COVID-19 patients when compared to mild/moderate cases (Table 2 and Figure 1A), while all others did not show differences between hospitalized and mild/moderate COVID-19 patients (Table 2). Spearman correlation analysis revealed that the plasma levels of MMP-1 and VEGF-A in hospitalized COVID-19 patients were positively correlated (Figure 1B), indicating that there are interactions between MMP-1 and VEGF-A in COVID-19. Thus, the highly elevated plasma levels of MMP-1 and VEGF-A were associated with the severity of COVID-19.

**Figure 1.**
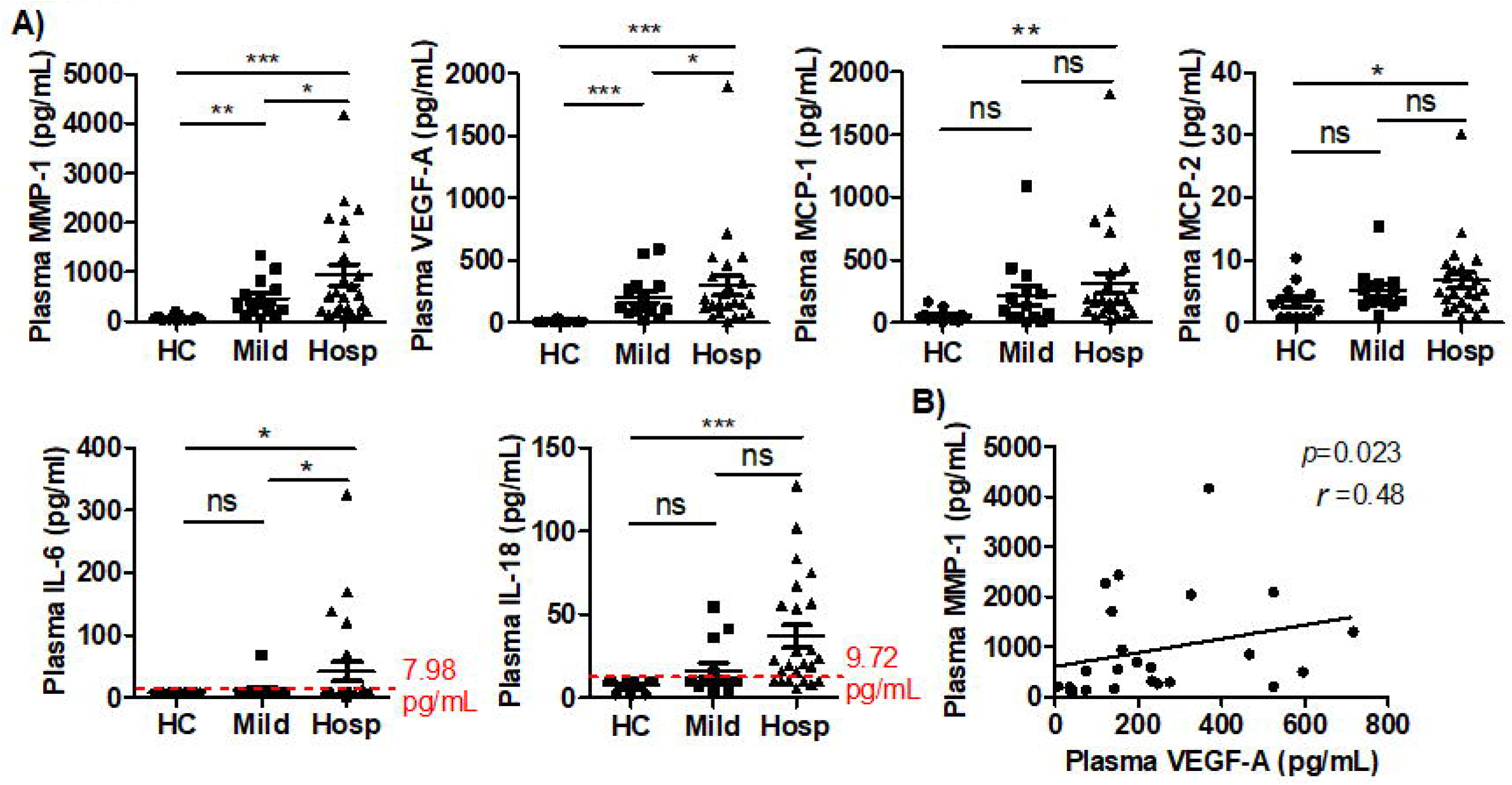
Plasma levels of inflammatory factors highly elevated in hospitalized COVID-19 patients. **A)** Scatter plots demonstrating the plasma levels of MMP-1, VEGF-A, MCP-1, MCP-2, IL-6, and IL-18 in hospitalized COVID-19 patients (n=24), mild/moderate COVID-19 patients (n=14), and HCs (n=13). Kruskal-Wallis test with Dunn’s correction for pairwise comparisons among hospitalized COVID-19 patients, mild/moderate COVID-9 patients, and HCs. **B)** Spearman correlation analysis between plasma levels of MMP-1 and VEGF-A in hospitalized COVID-19 patients. *r*, Spearman correlation coefficient. HC, healthy control; Mild, mild/moderate COVID-19 patients; Hosp, hospitalized COVID-19 patients. *\*p* < 0.05; *\*\*p* < 0.01; *\*\*\*p* < 0.001; ns, not significant; Horizontal lines represent the median.

MCP-1, MCP-2, IL-6, and IL-18 were the 4 analytes that were significantly increased in hospitalized patients, but not in mild/moderate cases, when compared to HCs (Table 2 and Figure 1A). However, plasma levels of IL-6 and IL-18 were elevated in some, but not all, patients with severe/critical COVID-19. As shown in Figure 1A, IL-6 was detected in 0 HCs (limit of detection ≥7.98 pg/mL), 1 mild/moderate case (67.8 pg/mL), and 7 hospitalized cases (13.8 pg/mL, 32.7 pg/mL, 68.9 pg/mL, 120.7 pg/mL, 138.2 pg/mL, 169.6 pg/mL, and 325.4 pg/mL). Similarly, IL-18 was detected in 0 HCs (limit of detection ≥9.72 pg/mL), 4 mild/moderate cases (15.3 – 54.4 pg/mL), and 18 hospitalized patients (15.7 – 127.3 pg/mL). Obviously, the levels of IL-6 and IL-8 are significantly less elevated in patients with critical COVID-19 than the values typically reported in patients with CSS such as septic shock with and without ARDS^24^.

Nineteen analytes were detected in all three groups of research subjects, but there were no differences between these groups (Table 2). Twenty-two analytes, primarily inflammatory cytokines such as IL-8 and TNF-α, were below the limit of detection in the majority of hospitalized cases, mild/moderate cases, and HCs (Table 2).

Our data support previous findings that suggested IL-6, IL-8, and TNF-α did not act as key inflammatory mediators for fatal manifestations in patients with severe or critical COVID-19^25^, and also question whether cytokine storm occurs in COVID-19.

### Enzymatic activity of MMP-1 and activation markers of ECs increased in the peripheral blood in hospitalized COVID-19 patients

MMP-1 and VEGF-A were two of the most unambiguously elevated inflammatory factors in hospitalized COVID-19 patients. Their plasma levels were positively correlated (Table 1 and Figure 1), suggesting that there are interactions between MMPs and ECs in COVID-19. To this end, we studied plasma MMP-1 enzymatic activity and the EC activation marker profiles. To date, only one MMP, MMP-9, has been analyzed in one pilot COVID-19 study^50^. This study measured plasma MMP-9 levels using an enzyme immunoassay, and showed that hospitalized COVID-19 patients had higher MMP-9 levels than HCs^50^. However, none of the MMPs have been studied for their enzymatic activity in COVID-19 patients. We analyzed enzymatic activity of MMP-1 in plasma samples from HCs and patients with mild/moderate or hospitalized COVID-19. We found that the enzymatic activity of MMP-1 was significantly increased in hospitalized COVID-19 patients when compared to HCs and those with mild/moderate COVID-19 (Figure 2A). Thus, both the levels and the enzymatic activity of MMP-1 were up-regulated in the peripheral blood of COVID-19 patients, particularly in patients with severe/critical COVID-19.

**Figure 2.**
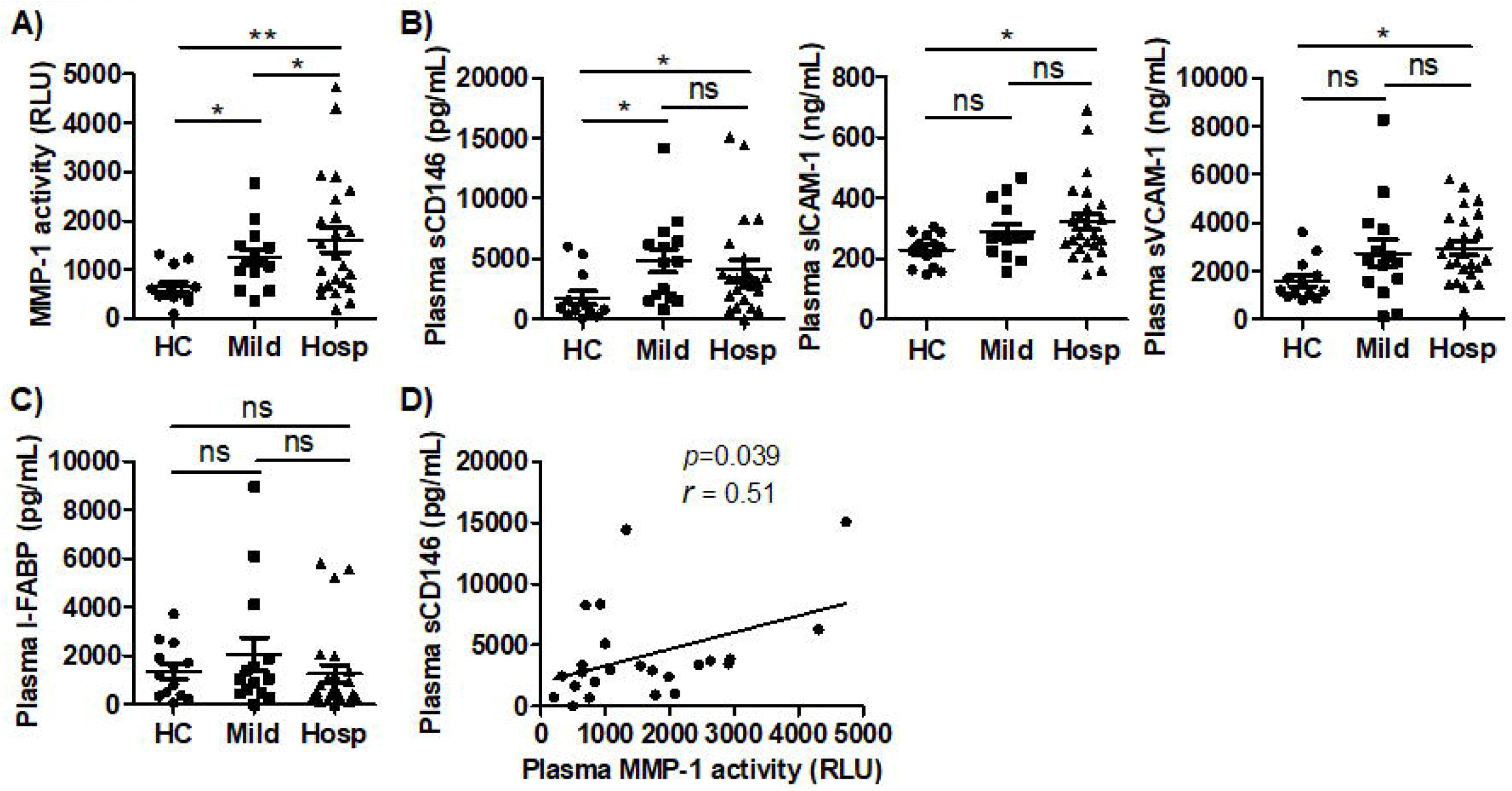
Enzymatic activity of MMP-1 and activation markers of ECs were elevated in the peripheral blood in hospitalized COVID-19 patients. **A), B), and C)** Scatter plots demonstrating enzymatic activity of MMP-1, levels of EC activation markers (sCD146, sICAM-1, and sVCAM-1), and level of intestinal epithelial injury marker (I-FABP), respectively, in the peripheral blood from hospitalized COVID-19 patients (n=24), mild/moderate COVID-19 patients (n=14), and HCs (n=13). Kruskal-Wallis test with Dunn’s correction for pairwise comparisons among HC, hospitalized COVID-19 patients, and mild/moderate COVID-9 patients. **B)** Spearman correlation analysis between plasma levels of MMP-1 and sCD146 in hospitalized COVID-19 patients. *r*, Spearman correlation coefficient. HC, healthy control; Mild, mild/moderate COVID-19 patients; Hosp, hospitalized COVID-19 patients. *\*p* < 0.05; *\*\*p* < 0.01; ns, not significant; Horizontal lines represent the median.

Next, we used ELISA assays to measure the levels of sCD146, sVCAM-1, and sICAM-1, surrogate markers of EC activation, in the plasma samples from hospitalized COVID-19 patients, mild/moderate cases, and HCs. As shown in Figure 2B, hospitalized COVID-19 patients had significantly higher plasma levels of sCD146, sICAM-1, and sVCAM-1, than HCs. Mild/moderate COVID-19 patients also had higher levels of sCD146, but neither sICAM-1 nor sVCAM-1, than HCs (Figure 2B). There were no differences in the plasma levels of sCD146, sICAM-1, or sVCAM-1 between hospitalized and mild/moderate COVID-19 patients (Figure 2B). As I-FABP is solely expressed in epithelial cells of the mucosal layer of the small intestine tissue and used as a plasma marker of intestinal epithelial injury^51,52^, I-FABP ELISA assay was also performed to study and compare the effects of COVID-19 on epithelial cells versus ECs. In contrast to EC activation markers, plasma I-FABP levels were no different among hospitalized COVID-19 patients, mild/moderate cases, and HCs (Figure 2C), suggesting that systemic ECs, not small intestine epithelial cells, are significantly affected by COVID-19.

Spearman correlation analysis was performed to identify any associations between plasma levels of active MMP-1 and EC activation markers. There was a positive correlation between plasma active MMP-1 and plasma levels of sCD146 (*r* = 0.51 and *p*<0.039) (Figure 2D). There were no correlations between the plasma levels of active MMP-1 and plasma levels of sICAM-1 or sVCAM-1 in hospitalized COVID-19 patients (data not shown).

Taken together, our data demonstrate that both plasma level and enzymatic activity of MMP-1 and plasma levels of EC activation markers are highly elevated and positively correlated in COVID-19 and their dysregulations are associated with the severity of COVID-19.

### Association of elevated MMP-1 and EC activation markers with demographics in hospitalized COVID-19 patients

Age is a well-known factor that influences the severity and fatality of COVID-19^53^. The risk for severe illness with COVID-19 increases with age, with older adults at higher risk^53^. Other demographic factors such as sex and race have been also linked with risk for severity of COVID-19 illness. We analyzed the relationship between demographic factors and circulating MMP-1 and EC activation markers in hospitalized COVID-19 patients. Twenty-four hospitalized COVID-19 patients were split into 2 age groups: 10 cases ≤ 55 years old and 14 cases > 55 years old. As shown in Figure 3A, plasma VEGF-A levels were significantly higher in hospitalized COVID-19 patients >55 years old when compared to cases ≤ 50 years old. Plasma levels of MMP-1, sICAM-1, sVCAM-1, active MMP-1 showed an increased trend in the older group (Figure 3A).

**Figure 3.**
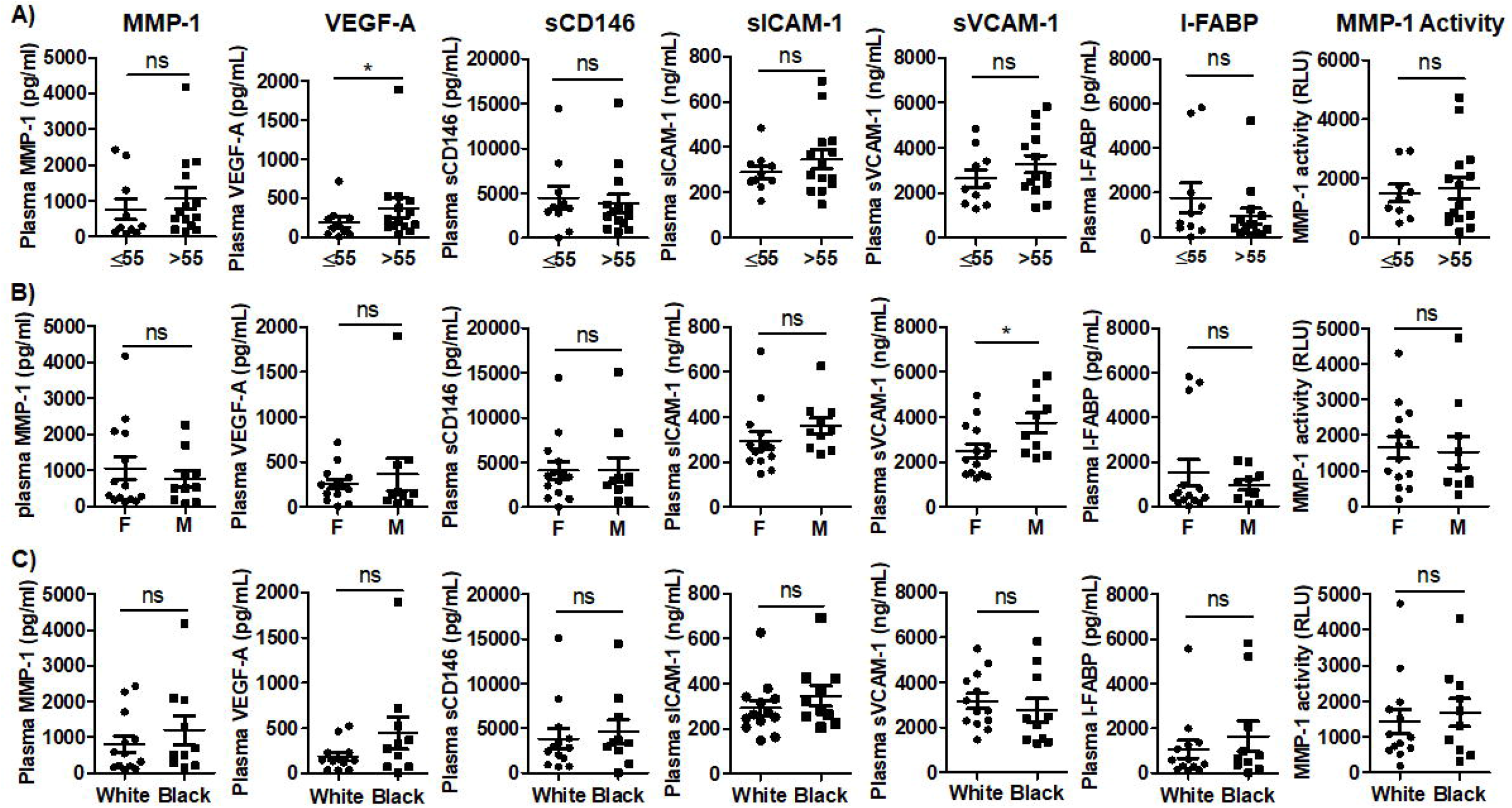
Relationship of MMP-1 and EC activation with demographics of hospitalized COVID-19 patients. Scatter plots demonstrating level of MMP-1, levels of EC activation markers (VEGF-A, sCD146, sICAM-1, and sVCAM-1), and level of intestinal epithelial injury marker (I-FABP), and enzymatic activity of MMP-1 in hospitalized COVID-19 patients. Comparison of these factors between hospitalized COVID-19 patients >55 years old (n=14) and ≤ 55 years old (n=10) **(A)**, female (n=14) and male (n=10) **(B)**, and African-Americans (n=10) and while Americans (n=13) **(C)**, respectively. All comparisons were conducted using Mann Whitney test. HC, healthy control; Mild, mild/moderate COVID-19 patients; Hosp, hospitalized COVID-19 patients; Black, African-American; White, Caucasian-American. *\*p* < 0.05; ns, not significant; Horizontal lines represent the median.

We also analyzed the associations of sex and race with MMP-1 and EC activation markers. As shown in Figure 3B and 3C, plasma sVCAM-1 levels were significantly higher in males (n=10) when compared to females (n=14) (Figure 3B). There were no differences in the plasma levels of MMP-1, active MMP-1, and EC activation markers in African-Americans (n=10) when compared to Caucasian Americans (n=13) (Figure 3C).

## Discussion

In the present study, we systematically profiled and compared 65 inflammatory cytokines/chemokines/growth factors in the plasma samples from hospitalized (severe/critical) COVID-19 patients, mild/moderate COVID-19 cases, and HCs. As shown in Table 2, hospitalized and mild/moderate COVID-19 patients had significantly higher plasma levels of 20 out of 65 factors analyzed when compared to HCs. These altered 20 factors include 10 chemokines (BLC/CXCL13, ENA-78/CXCL5, Eotaxin-3/CCL26, Fractalkine/CX3CL1, IP-10/CXCL10, I-TAC/CXCL11, MIG/CXCL9, MIP-1α/CCL3, MIP-1β/CCL4, and SDF-1α/CXCL12), 4 growth factors (HGF, MMP-1, SCF, and VEGF-A), 5 soluble receptors (APRIL, CD30, IL-2R/CD25, TNF-RII, and TRAIL/CD253), and 1 cytokine (MIF). In addition, four factors, MCP-1, MCP-2, IL-6, and IL-18, were significantly increased in hospitalized patients, but not in mild/moderate cases, when compared to HCs (Table 2, Figure 1). However, IL-6 and IL-18 were only elevated in some patients with severe/critical COVID-19, and their levels were significantly less elevated when compared to the values typically reported in patients with CSS such as septic shock with and without ARDS^24^. Thus, the vast majority of altered circulating inflammatory factors in COVID-19 patients were chemokines, growth factors, and soluble receptors, not inflammatory cytokines. Our results indicate that COVID-19 patients have distinct inflammatory profiles that are distinguished from the cytokine storms found in other human diseases^49,54,55^, and also question whether a cytokine storm occurs in COVID-19. The term “cytokine storm” was first coined to describe the excessive release of inflammatory cytokines by immune cells in graft-versus-host disease (GVHD)^49,54,55^. Since then, CSS has been reported in a wide range of other human diseases including cancer patients undergoing chimeric antigen receptor (CAR) T cell therapy^56,57^, pancreatitis^58^, multiple organ dysfunction syndrome^59^, multiple sclerosis^60^, sepsis^61^, and viral infections^21,62-68^. Indeed, CSS is a common complication of viral respiratory infections such as infection with influenza^62,63^, SARS-CoV^21,64,65^, and the Middle East respiratory syndrome coronavirus (MERS-CoV)^67,68^. In these CSS-linked diseases, several inflammatory cytokines including IL-6, IFN-γ, IL-1β, and TNF-α are highly elevated. In fact, elevated circulating levels of IL-6 are a hallmark of CSS in patients undergoing CAR T cell therapy^69^. In HCs, mean levels of circulating IL-6 have been reported to be <5 pg/mL^70^, which can be increased to >10,000 pg/mL in cancer patients on CAR T cell therapy^69^. The substantial elevation of IL-6 and its correlation with disease severity have resulted in IL-6 inhibitors such as sarilumab, siltuximab, and tocilizumab becoming therapeutic agents that can effectively treat CSS-associated diseases^69,71^. Circulating IL-6 levels are also elevated in COVID-19 patients and is cited as evidence of COVID-19 CCS^22,23,66,72-78^. However, as previously reported, elevated IL-6 levels in COVID-19 patients are minuscule compared to those found in individuals on CAR T cell therapy and other CSS-associated diseases^25^, suggesting that IL-6 does not act as a key inflammatory mediator for fatal manifestations in patients with severe or critical COVID-19^25^. In line with this finding, IL-6 inhibitors are not recommended to be used as therapeutic agents for the treatment of COVID-19 as noted by NIH guidelines^26^. Our results provide evidence that explains why efforts to combat cytokine storm have proven unsuccessful in severe COVID-19 patients. Our study implicates a chemokine storm, not a cytokine storm, occurs in COVID-19. This chemokine storm plays an important role in the pathogenesis of COVID-19 via recruitment of inflammatory cells to the lungs and other organs or tissues^1-5^. Thus, chemokines and chemokine receptors as therapeutic targets for COVID-19 treatment need to be studied.

As shown in Table 2 and Figure 1, MMP-1 and VEGF-A were two of the most unambiguously elevated inflammatory factors in hospitalized COVID-19 patients when compared to mild/moderate cases or HCs, while all other inflammatory factors did not show differences between hospitalized and mild/moderate COVID-19 patients. Spearman correlation analysis revealed that the plasma levels of MMP-1 and VEGF-A in hospitalized COVID-19 patients were positively correlated (Figure 1B), suggesting interactions between MMP-1 and VEGF-A in COVID-19. MMP-1 is an interstitial collagenase capable of degrading collagen types I, II, and III, and plays a critical role in vascular remodeling and vascular diseases^79,80^. In addition, MMP-1 acts as a potent agonist for protease-activated receptor-1 (PAR1)^81^, a G protein-coupled protease-activated receptor, on the surface of a variety of cell types such as ECs^81-83^, platelets^81,84,85^, and macrophages^86,87^. MMP-1/PAR1 signaling plays a paramount role in EC function^81-83^. Hyperactivation of MMP-1/PAR1 signaling in ECs leads to a decreased endothelial barrier function and promotes adhesion of inflammatory cells to ECs^88-90^. MMP-1/PAR1 signaling can increase expression of VEGF receptor-2 (VEGFR2)^91^, the main receptor for VEGF-A^92^, on ECs. The increased VEGFR2 binds VEGF-A, resulting in increased activation of ECs. We found that the levels of MMP-1, VEGF-A, and MMP-1 enzymatic activity were significantly elevated in the peripheral blood in hospitalized (severe/critical) COVID-19 patients compared to mild/moderate cases or HCs (Figure 2), suggesting that MMP-1/PAR1/VEGFR2/VEGF-A signaling may be in a hyperactivation state in ECs in COVID-19 patients.

In addition to VEGF-A, other EC activation markers including sCD146, sICAM-1, and sVACM-1 were highly elevated in COVID-19. In contrast, plasma I-FABP levels were no different between COVID-19 patients and HCs (Figure 2C). These findings indicate that systemic ECs, not small intestine epithelial cells, are significantly affected in COVID-19. Taken together, our results imply that excessive MMPs are strongly related to the severity of COVID-19, likely playing a central role in widespread damage to the vascular endothelium. In humans, there are 23 related, but distinct, MMPs. Thus, there is an urgent need to study the profile, expression, and activity of MMPs in SARS-CoV-2 infection and the role of excessive MMPs in the pulmonary and systemic endothelialitis and vascular injury in COVID-19 patients. A better understanding of MMP profiles and their pathological roles will provide insights into application of therapeutic MMP inhibitors for COVID-19 treatment.

Age is a well-known factor that influences the severity and fatality of COVID-19^53^. The risk for severe illness with COVID-19 increases with age, with older adults at highest risk^53^. Other demographic factors such as gender and race have been also linked with risk for the severity of COVID-19 illness. We found that excessive MMPs and/or circulating markers of EC activation increased with age or were higher in hospitalized male COVID-19 patients (Figure 3), but not affected by race (Figure 3). However, we realize that our sample size of hospitalized COVID-19 patients, after splitting into 2 demographic groups, is too small to make statistical inferences.

## Data Availability

We would wish to make our results available to the community of scientists interested in COVID-19. We would also welcome collaboration with others who could make use of the strategies and reagents developed in this project. As researchers and educators, we would also make our findings available to the public.

https://medicine.iu.edu/faculty/6507/yu-andy

## Acknowledgments

This work was partially funded by NIAAA grant (UH2AA026218 to Q.Y.) and the grant (OPP1035237 to Q.Y.) from the Bill & Melinda Gates Foundation. This publication was made possible, in part, with support from the Indiana Biobank and the Indiana Clinical and Translational Sciences Institute (CTSI) funded by the National Institutes of Health (UL1TR002529), National Center for Advancing Translational Sciences, Clinical and Translational Sciences Award. The content is solely the responsibility of the authors and does not necessarily represent the official views of the National Institutes of Health.

Authors have no conflict of interest to declare.

## Abbreviations

ACE2: angiotensin-converting enzyme 2
ALC: absolute lymphocyte count
APMA: amino-phenyl mercuric acetate
ARDS: acute respiratory distress syndrome
CAR: chimeric antigen receptor
COVID-19: coronavirus disease 2019
CEC: circulating endothelial cell
CRP: C-reactive protein
CSS: cytokine storm syndrome
EC: endothelial cell
ESR: erythrocyte sedimentation rate
GVHD: graft-versus-host disease
HC: healthy controls
HGF: hepatocyte growth factor
ICAM-1: intercellular adhesion molecule 1
I-FABP: intestinal fatty acid binding protein
ICU: intensive care unit
DIC: disseminated intravascular coagulation
MERS-CoV: East respiratory syndrome coronavirus
MIF: macrophage migration inhibitory factor
MMP-1: matrix metalloproteinase-1
NRP1: neuropilin-1
NT-proBNP: N-terminal (NT)-pro hormone B-type natriuretic peptide
PAR1: protease-activated receptor-1
PBS: phosphate buffered saline
PBMC: peripheral blood mononuclear cell
RFU: relative fluorescence unit
SARS-CoV-2: severe acute respiratory syndrome coronavirus 2
SD: standard deviation
SEM: standard error of the mean
TMPRSS2: transmembrane protease serine 2
VCAM-1: vascular cell adhesion molecule 1
VEGF-A: vascular endothelial growth factor A.

